# Early medical abortion by telemedicine in the United Kingdom: A cost-effectiveness analysis

**DOI:** 10.1101/2021.02.26.21252518

**Authors:** James E Hawkins, Anna Glasier, Stephen Hall, Lesley Regan, on behalf of the RCOG Telemedicine cost-effectiveness working group, Homerton University Hospital Foundation Trust, University Hospitals Bristol NHS Foundation Trust, Aneurin Bevan University Health Board

## Abstract

**Objective:** To determine the potential cost savings resulting from the introduction of routine early medical abortion at home by telemedicine in the United Kingdom.

**Design:** A cost-effectiveness analysis Setting: United Kingdom

**Population:** Women in 2020 undergoing early medical abortion provided by three independent abortion providers and two NHS abortion clinics.

**Methods:** Computation of the costs of each abortion procedure and of managing failed or incomplete abortion and haemorrhage requiring blood transfusion.

**Outcome measures:** Cost savings

**Results:** Overall estimated cost savings are £15.80 per abortion undertaken by independent abortion providers representing a saving to the NHS of over £3 million per year. Limited data from NHS services resulted in an estimated average saving of £188.84 per abortion.

**Conclusions:** Were telemedicine EMA to become routine, an increase in the number of women eligible for medical rather than surgical abortion, and a reduction in adverse events resulting from earlier abortion could result in significant cost-savings.

**Tweetable Abstract:** Early medical abortion at home using telemedicine could save the NHS £3 million per year

## Introduction

The testing of a healthcare intervention should answer three questions about its efficacy, effectiveness and efficiency ^1^. The gold standard for testing efficacy is a randomised controlled trial (RCT). Meticulously designed and conducted, RCTs show whether an intervention *can* work under ideal conditions. But delivery of healthcare in the community is often imperfect, and effectiveness studies are required to show whether a promising intervention works in typical circumstances. To show that the intervention *does* work, a community intervention study is necessary. Efficiency studies are designed to determine whether the intervention is cost-effective, whether it is ‘worth it’.

Abortion is one of the commonest gynaecological interventions globally. Over 207,000 abortions were undertaken in England and Wales in 2019, all of them, at least for part of the procedure, in a clinical setting ^2^. Numerous efficacy studies have shown that early medical abortion (EMA) can be done safely and effectively at home without any face-to-face interaction with a healthcare provider ^3,4^. Very recently a community intervention study has shown that EMA undertaken by telemedicine at home in the UK *does* work, that it is at least as safe and effective as EMA undertaken in a clinical setting ^5^.

In March 2020, in response to the Covid-19 pandemic, three large independent abortion providers in England and Wales began offering telemedicine EMA (TM-EMA) to all eligible women. Telephone consultations were used to assess the woman and inform them about the abortion procedure, ultrasound examination was only done when there was doubt about the gestation or possible ectopic pregnancy, and both mifepristone and misoprostol were sent to eligible women who took both medicines at home ^4^. An analysis of data available before and after March 2020 showed no difference between TM-EMA and the service-based care provided previously in either the rate of complete abortion or the overall incidence of adverse events. The mean waiting time for abortion was shorter by 4.2 days in the telemedicine cohort, and many more procedures were provided before six weeks gestation ^5^.

In this paper we have used the data from the community intervention study ^5^ to try to answer the third question about the abortion at home, is it worth it - is TM-EMA cost-effective and are there potential cost savings compared with traditional service-based provision? We hypothesised that a reduction in the waiting time between initial presentation and the abortion procedure, thereby reducing the gestation at which abortion was done, would enable more women to choose TM-EMA and reduce the number of the more expensive surgical procedures which are undertaken in a clinical setting. We further hypothesised that a higher rate of EMA, together with an increase in the number of abortions undertaken at very early gestations, should lead to a reduction in the incidence and severity of adverse events requiring further intervention at additional cost.

## Methods

We used the same approach as that described in the NICE Guideline on Abortion Care 2019 ^6^. In the primary analysis we used information from independent abortion providers to compare data from all women having an EMA with traditional management during January 1^st^ - March 1^st^ 2020, with data from all women undergoing TM-EMA between April 6^th^ and June 30^th^ 2020.

It was not possible to collect a similar data set from the large number of much smaller NHS services providing different models of abortion care throughout the country and so not possible to compute estimated savings for the 26% of abortions done in NHS services. In order to determine whether the switch to TM-EMA in NHS settings would likely result in similar cost savings, we analysed separately data sets from two NHS abortion services ((University Hospitals Bristol NHS Foundation Trust in Bristol and Aneurin Bevan University Health Board in Wales) who provided activity data on abortion both before (1^st^ January-29^th^ February 2020) and after (1^st^ May to 30^th^ June 2020) introducing TM-EMA.

Finally, we used national data from The Abortion Statistics for England and Wales: 2019 to determine the annual number of abortions and the number that would be eligible for TM-EMA over the period of one year ^2^. In 2019, a total of 207,105 abortions were performed before 24 weeks gestation in England and Wales, 82.5% of them before the end of the 9^th^ week of pregnancy and these would have been potentially eligible for TM-EMA. In 2019, 75% of abortions in England and Wales were performed in the independent sector and this percentage was used for computation of overall savings made from TM-EMA done in that sector. Since the number of abortions has been more than 190,000 per year since 2009 it was felt that the 2019 figures would likely represent the annual number of abortions in the future.

Costs of the abortion procedures in the analysis of the independent sector data were calculated from tariff prices already in place in England and Wales. These contracts, agreed with NHS Clinical Commissioning Groups (CCG), give a fixed price for providing an abortion, based mainly on gestational age and type of procedure. Remunerations were identical regardless of whether the abortion was TM-EMA or EMA performed in a clinical setting. Agreed contract prices may differ between providers but they are confidential and therefore have not been reported in a disaggregated form. A mean cost per abortion across all gestational weeks and types of procedure was therefore calculated (table 1).

**Table 1:**
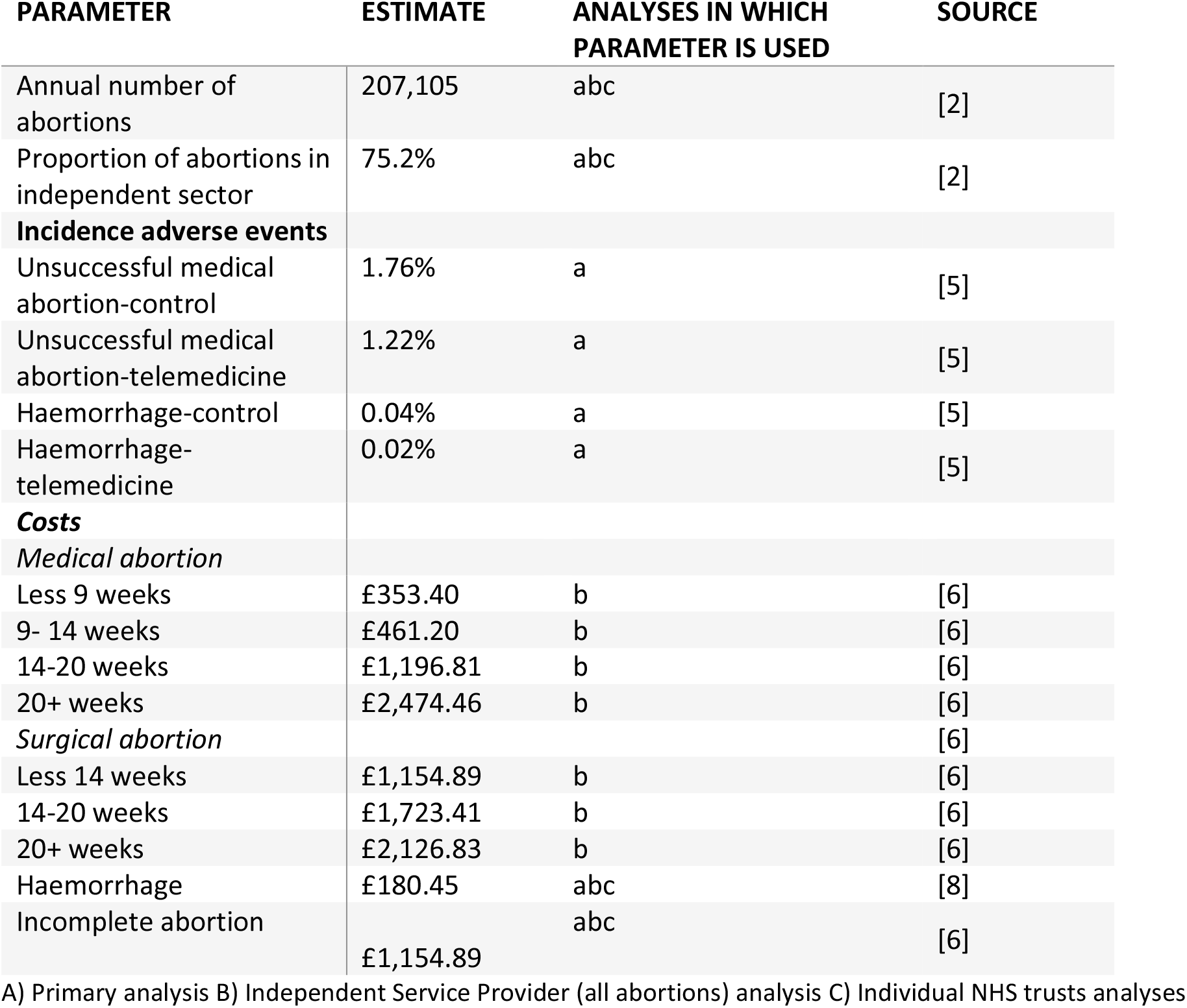
Parameter estimates and statistical distributions used in the economic model

Remuneration data were obtained from the largest two national abortion providers the British Pregnancy Advisory Service (BPAS) and MSI Reproductive Choices (MSUK) for the period 1st January to 29^th^ February 2020 (traditional care costs) and 1^st^ May to 30^th^ June 2020 (for MSUK) and 8^th^ April to 8^th^ June (for BPAS) (TM-EMA costs). The difference in dates for the TM-EMA data was due to different data collection methods between the two organisations and was considered unlikely to affect any overall cost estimates. A difference in cost between the two periods was calculated by dividing the total remuneration from CCG contracts by the total number of abortions. Where contract prices had changed between the two periods, the traditional care costs were calculated by multiplying the number of procedures by the TM-EMA period tariff.

For the analysis of data from NHS services the cost of abortion procedures were taken from NHS Improvement ^7^ and are also shown in Table 1. Tariffs differed for medical and surgical abortions according to gestation, and for elective, non-elective long stay, non-elective short stay, day case and outpatient settings. A weighted mean cost for medical and surgical abortions was calculated for each gestational period reported based on the reported activity level. Compared with the contracts agreed by the CCGs with independent abortion providers, NHS tariff costs are higher and have larger cost differences between different methods and gestations. Comparably, small changes in the numbers of surgical abortions undertaken after ten weeks gestation to early medical abortion have a large impact on cost.

Agreeing a cost for adverse events was complicated. In the independent sector most of the common adverse events are dealt with by the abortion provider and are included in the tariff received from the CCGs (despite imposing additional costs on the provider). Serious adverse events requiring visits to emergency departments and/or hospitalisation are rare. The overall incidence of ectopic pregnancy was equivalent in both cohorts^5^, 39 cases (0.2%) in the traditional, and 49 (0.2%) in the telemedicine pathway so we did not include costs for managing ectopic pregnancy. In the community intervention study ^5^ no-one in either group (traditional care or TM-EMA) required major surgery or intravenous antibiotics for infection. Haemorrhage requiring transfusion and failed/incomplete abortion requiring surgical intervention to manage continuing pregnancy or retained products of conception (POC) were included in the model as they were associated with additional costs to the NHS not just to the independent abortion provider (Table2). The cost of a haemorrhage was taken from the costings for one blood transfusion from the health economic model for the NICE Guideline NG24: Blood Transfusions^8^ and inflated to the 2018/19 price using the hospital & community health services (HCHS) index ^9^ and estimated at a cost of £198.74 per transfusion. It was assumed that any haemorrhage would require only one transfusion. The cost of surgical intervention for continuing pregnancy or retained POC were assumed to be equal to that of a surgical abortion between 0-14 weeks as calculated for the NHS cost (Table 1). Data on adverse events for abortions undertaken in NHS services was not available.

**Table 2:** Estimated total cost savings to the CCGs from the introduction of a telemedicine model of abortion - primary analysis

|  | **COST SAVING PER**<br>**ABORTION TELEMEDICINE**<br>**MODEL** | **SAVING TO NHS FROM**<br>**INDEPENDENT SECTOR**<br>**ONLY** | **SAVING TO NHS PER**<br>**YEAR†** |
| --- | --- | --- | --- |
| **A) ABORTION**<br>**PROCEDURE COST** | £9.60 | £1,494,087 | £1,988,163 |
| **B) ADVERSE EVENT-**<br>**INCOMPLETE**<br>**ABORTION** | £6.18 (-£8.09, £8.08) ‡ | - | £1,279,444 (-£1,675,138,<br>1,672,724) |
| **C) ADVERSE EVENT-**<br>**HAEMORRHAGE** | £0.02 (£0.02, 0.02) | - | £4,768(-£4765, £4771) |
| **TOTAL (A+B+C)** | £15.80 |  | £3,272,375 |

Abortion procedure cost was calculated by multiplying the cost saving per abortion with all abortions performed in England and Wales. For all other outcomes, costs were multiplied by the total number of early medical abortions.
‡The confidence intervals for this outcome are calculated using the adjusted p-value from Aiken 2021 <sup>5</sup>.

95% confidence intervals were estimated for total costs based upon the adjusted p-values reported for the effectiveness outcomes. Contract prices and NHS tariffs were assumed to be fixed and therefore confidence intervals could not be calculated for some components of total costs.

Set-up costs were not considered in the cost-effectiveness analysis.

Only costs being borne by the NHS and ultimately by the UK Treasury were included in the economic analysis. We did not include costs to the women themselves nor did we attempt to estimate any changes in quality of life.

## Results

### Abortions undertaken by the independent providers

MSUK and BPAS provided financial data for a total of 26,405 early medical abortions for the traditional care period and 26,430 EMA’s for the TM-EMA period. Some 76% of procedures performed in the traditional care period were early medical abortions compared to 86% in the post period. This resulted in a reduction in average tariff paid by CCGs of £9.60 per abortion. (Table 2) If this saving were realised for all abortions performed in England and Wales regardless of provider, it would represent a total cost saving of £2 million per year. If it were realised only for the abortions undertaken in the independent sector a cost saving of £1.5million per year would be realised.

The reduction in serious adverse events (as a result of more abortions being done at earlier gestation) adds to the cost savings. There were 389 (1.8%) and 366 (1.2%) failed/incomplete abortions in the traditional care and TM-EMA periods respectively, a difference of less than 1 percentage point. The additional cost saving from this reduction in adverse events was £6.18 for every abortion. Haemorrhage requiring blood transfusion reduced costs by only £0.02 per abortion.

Overall estimated cost savings are £15.80 per abortion representing a saving to the NHS of over £3 million per year. The saving on the cost of the procedure itself represents almost two thirds of the overall cost saving and is the only area of cost saving for which the 95% confidence intervals did not go below zero where the telemedicine model becomes cost increasing. Over a third of the total cost savings came from a reduction in failed/incomplete abortions although the 95% confidence intervals pass zero where the telemedicine model is more expensive. The cost of managing haemorrhage made up less than 1% of all costs.

The independent service providers (BPAS, MSI Choices, and NUPAS) also provided data on all the abortions undertaken from January to July 2020 (both before and after the introduction of telemedicine) including surgical abortions and medical abortions at higher gestation - a total of 27,865 abortions for the period before introduction of TM-EMA and 33,809 for the period after TM-EMA. Adverse events were not included in this dataset. These data indicated a slightly greater shift from surgical to medical abortion than the data used in the primary analysis – with 76% of abortions in the pre period and 89% in the post period being EMAs (Table 3). This shift from more costly, later, surgical procedures to EMA has resulted in a similar estimated cost saving to the primary analysis – of £9.95 per procedure, or an overall annual NHS saving of just over £2 million (Table 3).

**Table 3:**
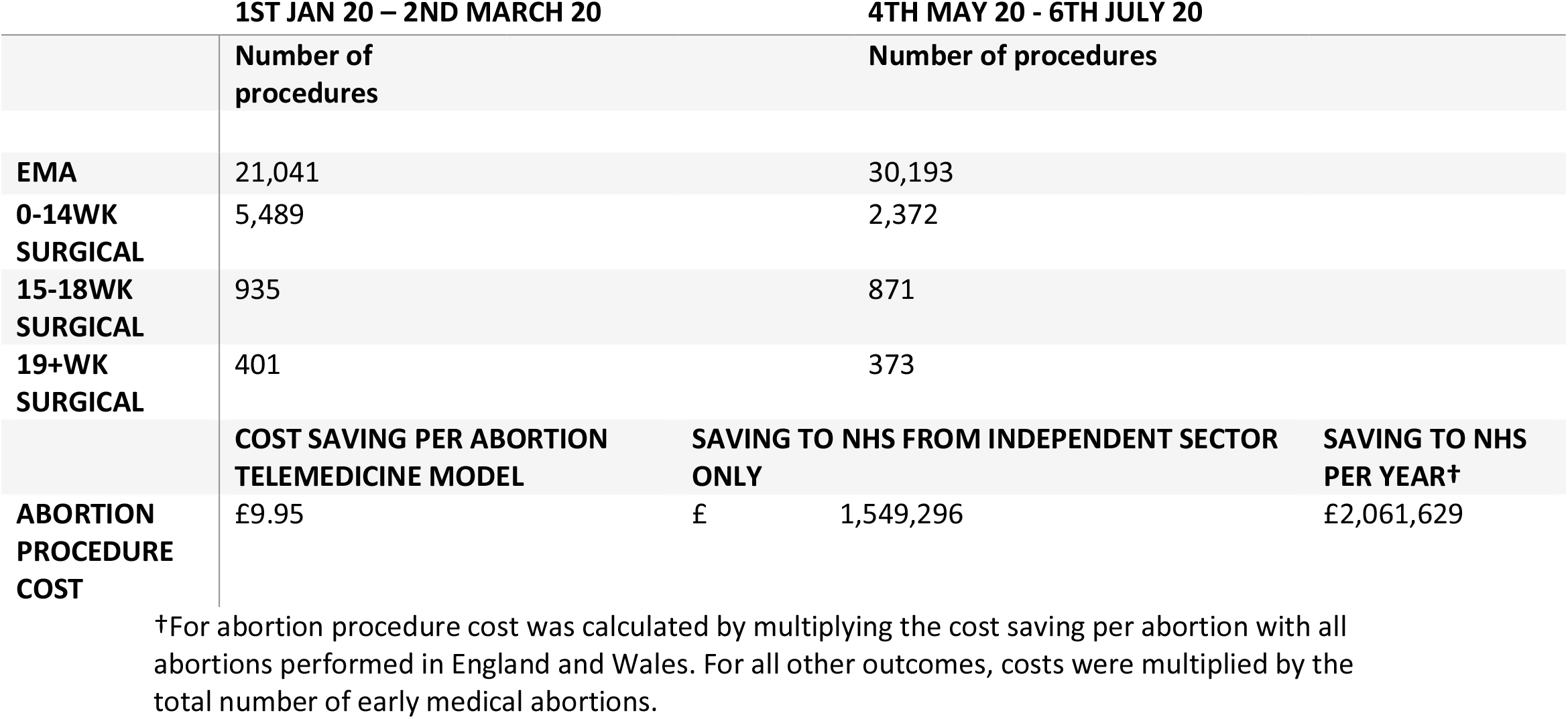
Independent Service Provider (all abortions) analysis

## NHS abortion services

Aneurin Bevan University Health Board, Wales provided 309 abortions in the period of TM-EMA, compared to 327 abortions undertaken in the period before the change in the law. Between these two time periods, there was a sizeable shift towards both TM-EMA and to abortions undertaken at earlier gestation. Before the approval of TM-EMA 69% of abortions were undertaken by EMA, compared to 91% after the introduction of TM-EMA. The most sizeable change was in the number of surgical abortions done up to 14 weeks gestation which declined from 91 procedures to 21. The shift in gestation and method resulted in an estimated average saving of £188.84 per abortion.

University Hospitals Bristol NHS Foundation Trust reported an increase in abortion numbers from 210 before the switch to TM-EMA to 248 afterwards. As with other services, there was a sizeable shift from surgical abortion (provided by this service only up to 14 weeks) to EMA. EMA accounted for 75% of abortions before the introduction of TM-EMA, compared to 98% after its introduction. As a result, this service saw an estimated average cost saving of £182.89 per abortion.

## Discussion

The main strength of this analysis is that only three provider organisations were involved in the community intervention study, all undertaking early medical abortion using a very similar approach and producing a very large data set. Data coming from NHS abortion providers are much less robust but have been included in order to enable extrapolation of the findings to all abortion providers in the UK. Data on adverse events associated with the abortion procedure which add to the overall cost were not available from NHS providers, but we have no reason to suspect that the incidence of these events would be any different. A further strength is the use of an economic model which was developed and already tested using national UK data ^6^. Data on the cost of managing adverse events is not easy to compute. As stated earlier, the majority of adverse events which occur during abortion procedures undertaken by the independent providers are managed by those providers either at the time of the first procedure or at a review visit (depending on the nature of the adverse event). Data from BPAS suggest that around 83% of incomplete abortions are managed by the independent providers once the diagnosis is made and just 17% of cases require hospital treatment e.g. for blood transfusion. [BPAS personal communication] Likewise, 86% of failed abortions resulting in ongoing pregnancies requiring a repeat abortion procedure were managed by BPAS themselves and only 14% were managed in an NHS hospital. While managing such adverse events incurs additional costs for the independent providers the CCG tariff takes no account of this. So, the costs of the adverse events in this analysis may be marginally overestimated.

While the amount of data from NHS abortion services was very limited, it is worth noting the very much larger cost saving involved compared with abortions undertaken by the independent providers. A number of possible reasons account for this difference. Firstly, in England and Wales most NHS services only undertake abortions for women who have existing medical conditions which contraindicate care in a setting with limited clinical facilities or where there might be a risk of serious complications, a morbidly obese woman for example, who for some reason requires general anaesthesia. These cases are much more complex and hence more expensive. Secondly, independent providers undertake large numbers of abortions and can apply substantial economies of scale not available to individual NHS hospitals some of whom are doing only a handful of abortions each month. Most of the large cost saving in the NHS results from the shift from surgical to medical and the large price differential between them. When comparing an EMA to a 0-14week surgical abortion the price difference in the independent providers contract costs is less than £100 whilst it is over £800 in the NHS.

The cost-effectiveness analysis shows that retaining the model of telemedicine for the management of early medical abortion would result in substantial cost savings to the UK NHS of between £1.5 and £3 million every year. The TM-EMA procedure is simple, has been endorsed by the national professional bodies including the Royal College of Obstetricians and Gynaecologists, Faculty of Sexual and Reproductive Healthcare, Royal College of Midwives, British Society of Abortion Care Providers) ^10^ and could be adopted by all NHS abortion providers as well as providers in other parts of the world. With additional evidence from the intervention study, it should be possible to refine the telemedicine model further, for example by reviewing the effectiveness of the algorithm for determining the need for pre-abortion ultrasound or the 10-week gestational age limit for eligibility in England and Wales (which was not imposed in Scotland). Such refinements would be likely to increase the number of women eligible for TM-EMA and thereby increase the cost-savings.

During the coronavirus pandemic, the NHS has struggled to provide care for conditions unrelated to Covid-19. Waiting lists continue to grow as public health challenges mount. Yet this innovative model of delivering early medical abortion has shortened waiting times, reduced the average gestation at the time of the abortion procedure, improved safety and efficacy and provided substantial cost savings.

If TM-EMA becomes standard care in the UK NHS and if these cost-savings are realised it will, of course, be up to the government as to how such savings could be used. Providing improved access to contraception is one way in which these savings can be multiplied to further enhance women’s health services and potentially to reduce the number of unintended pregnancies and abortions. Public Health England reports that contraception is the single most cost-effective intervention in healthcare, and that for each £1 spent on contraception, it is estimated that £9 is saved in downstream costs^11^.

National guidance recommends that women should be offered a choice of abortion procedure^6^. In a sub-sample of women undergoing TM-EMA, 95% were satisfied or very satisfied with the procedure and 80% would choose TM-EMA in the event of needing another abortion, just 13% would have preferred in-person care ^5^. Fear of attending clinical services during the coronavirus pandemic may have led to more women being happy to accept telemedicine than previously, on the other hand were TM-EMA to become the norm in the future, familiarity (among both users and providers) may make it even more acceptable.

Costs to the services for setting up TM-EMA were not included in the analysis. It was assumed that all clinics would already have appropriate phone/video conferencing facilities, but some produced additional information materials (often in video format).

In this analysis we have not taken into account costs to the women undergoing abortion nor quality of life aspects. A systematic review of TM-EMA ^4^ identified out-of-pocket costs for women including travel, childcare and time off work – most, if not all, of which could be averted with telemedicine adding to the cost savings to society as a whole. For those women who choose to attend a clinic for abortion we would hope to see a better experience due to reduced pressure on these services. Policymakers should consider the quality-of-life savings for women. Women eligible for TM-EMA encounter fewer logistical barriers associated with potentially multiple trips to a clinic to attend appointments, often during normal working hours. More research should be undertaken in this area to consider the potential extent of these financial savings to both women and the wider economy.

We should consider, together with evidence on clinical outcomes, how this analysis can help to define a new abortion care pathway which can be delivered safely and efficiently worldwide. Telemedical models have advantage where access to infrastructure and resources for health service delivery are limited and women typically often travel long distances to access care.

In conclusion, introduction of TM-EMA as routine in the UK could result in cost-savings of up to £3 million per year. Looking to the future the telemedicine model of abortion without the need for routine ultrasound and with the procedure itself undertaken at home by women administering the drugs to themselves would make early medical abortion an intervention which could easily be administered from a general practice setting in the UK. Medical abortion could join the growing list of self-care interventions which increase choice, accessibility, and affordability, as well as opportunities for individuals to make informed decisions regarding their health and healthcare.

## Data Availability

Data openly available in a public repository that issues datasets with DOIs. Other data available on request.

https://obgyn.onlinelibrary.wiley.com/doi/10.1111/1471-0528.16668

